# Structural changes to primary visual cortex in the congenital absence of cone input in achromatopsia

**DOI:** 10.1101/2021.07.19.21260427

**Authors:** Barbara Molz, Anne Herbik, Heidi A. Baseler, Pieter B. de Best, Richard Vernon, Noa Raz, Andre Gouws, Khazar Ahmadi, Rebecca Lowndes, Rebecca J. McLean, Irene Gottlob, Susanne Kohl, Lars Choritz, John Maguire, Martin Kanowski, Barbara Käsmann-Kellner, Ilse Wieland, Eyal Banin, Netta Levin, Michael B. Hoffmann, Antony B. Morland

## Abstract

Autosomal recessive Achromatopsia (ACHM) is a rare inherited disorder associated with dysfunctional cone photoreceptors resulting in a congenital absence of cone input to visual cortex. This might lead to distinct changes in cortical architecture with a negative impact on the success of gene augmentation therapies. To investigate the status of the visual cortex in these patients, we performed a multi-centre study focusing on the cortical structure of regions that normally receive predominantly cone input. Using high-resolution T1-weighted MRI scans and surface-based morphometry, we compared cortical thickness, surface area and grey matter volume in foveal, parafoveal and paracentral representations of primary visual cortex in 15 individuals with ACHM and 42 normally sighted, healthy controls (HC). In ACHM, surface area was reduced in all tested representations, while thickening of the cortex was found highly localized to the most central representation. These results were comparable to more widespread changes in brain structure reported in congenitally blind individuals, suggesting similar developmental processes, i.e., irrespective of the underlying cause and extent of vision loss. Our findings indicate that there may be an optimum time window for gene therapy to counteract developmental cortical changes related to the absence of sensory input.

**Highlights:**

- We assessed cortical anatomy of a large cohort of patients with genetically confirmed cone photoreceptor dysfunction (achromatopsia) using surface-based morphometry.
- We found widespread reduction in cortical surface area across foveal, parafoveal and paracentral proportions of primary visual cortex in participants with achromatopsia.
- highly localized cortical thickening in participants with achromatopsia at the region of visual cortex that lacked inputs from the retinal region occupied solely by cones.
- Further evidence that the visual cortex is unlikely to take on normal properties if vision were restored after the developmental plastic period.
- Early intervention is preferable when considering vision restoration treatment in achromatopsia.

## Introduction

With the rise of gene therapeutic interventions to treat congenital ophthalmological diseases, assessing the structural integrity of visual cortex is critical as successful visual rehabilitation will ultimately rely on intact brain function. Achromatopsia (ACHM), a rare (1:30,000) monogenic, autosomal recessive visual disorder that affects signal transduction in cone photoreceptors (Aboshiha et al., 2016; Michalakis et al., 2017; Zobor et al., 2015), is currently the target of gene-addition therapy in a number of clinical trials (NCT03758404, NCT03001310, NCT03278873, NCT02935517, NCT02599922, NCT02610582, (Fischer et al., 2020)).

As cones are the only photoreceptors to occupy the central foveal region of the retina, patients with ACHM have a central scotoma and therefore reduced visual acuity from birth, along with an absence of colour vision, photophobia, hemeralopia and nystagmus (Haegerstrom-Portnoy et al., 1996; Remmer et al., 2015; Zobor et al., 2015). While the overall stationary or very slow progressing disease phenotype (Zobor et al., 2015) would generally be favourable for therapeutic intervention at any point during the lifespan in ACHM (Hirji et al., 2018) the way in which cortical structure develops during extended periods of cortical deprivation of cone inputs has not yet been assessed. It is plausible that structural remodelling of the visual cortex due to lack of input may limit the processing of restored signals from the eye. Structural differences in the visual cortex have been reported in congenital visual disorders such as congenital anophthalmia, congenital glaucoma, albinism and retinitis pigmentosa (Bridge et al., 2014, 2009; Jiang et al., 2009; Park et al., 2009). Several studies using voxel-based morphometry (VBM) reported reduced grey matter volume in early blind individuals (Noppeney et al., 2005; Pan et al., 2007; Park et al., 2009; Ptito et al., 2008; Von Dem Hagen et al., 2005). Further studies applied a surface-based approach and linked the volumetric reduction to a decrease in surface area (Aguirre et al., 2016; Jiang et al., 2009; Park et al., 2009). Reductions in cortical volume and surface area were attributed in part to the loss of input to visual cortex early in life (Jiang et al., 2009; Park et al., 2009; Ptito et al., 2008). Independently, and perhaps counterintuitively however, a number of studies also found increased cortical thickness, e.g. in Leber congenital amaurosis (Aguirre et al., 2017), congenital anophthalmia (Bridge et al., 2009), Leber’s hereditary optic neuropathy (d’Almeida et al., 2013) and other forms of congenital blindness (Aguirre et al., 2016; Anurova et al., 2015; Jiang et al., 2009; Park et al., 2009). Although the exact mechanisms behind these cortical changes have not yet been proven, there is evidence that they may indicate some form of neuronal degeneration or reorganization which could limit the outcome of vision restoration treatments (Guerreiro et al., 2016, 2015; Lemos et al., 2016; Prins et al., 2016a).

Understanding whether ACHM specifically affects the morphology of brain regions that process central vision is therefore a crucial factor in evaluating the potential success of gene augmentation therapies in this patient population. We hypothesise that individuals with ACHM will show distinct changes in cortical anatomy similar to those found in early blind individuals, but distinctly in central visual field representations of primary visual cortex that have been deprived of normal cone input throughout development.

## Materials and Methods

### Participants

Data used in this study were collected as part of a multicentre project at three scanner sites (University of York, UK (“UY”), Hadassah Medical Center, Jerusalem, IL (“HMC”), University of Magdeburg, DE (“UM”)). High resolution structural scans from 42 participants (mean ± SD age, 30.29 ± 9.72; 19 males) with normal or corrected-to-normal vision (HC) and 15 participants with genetically confirmed ACHM (biallelic *CNGA3* or *CNGB3* mutations as specified in suppl. Table S1; (age mean ± SD, 36.2 ± 10.26; 9 males) and electroretinographically confirmed absence of cone-function were utilised in this study. Experimental protocols received approval from the site-specific ethics committees and were in accordance with the Declaration of Helsinki.

### Data acquisition

#### York

A single, high resolution, anatomical, T1-weighted scan (TR, 2500ms; TE, 2.26ms; TI, 900 ms; voxel size, 1×1×1mm^3^; flip angle, 7°; matrix size, 256×256×176) was acquired using a 64-channel head coil on a SIEMENS MAGNETOM Prisma 3T scanner at the York Neuroimaging Centre (YNiC).

#### Jerusalem

A single, high resolution, anatomical, T1-weighted scan (TR, 2300ms; TE, 2.98ms; TI, 900 ms; voxel size, 1×1×1mm^3^; flip angle, 9°; matrix size, 256×256×160) was acquired using a 32-channel head coil on a SIEMENS MAGNETOM Skyra 3T scanner at the Edmond & Lily Safra Center for Brain Sciences, Hebrew University of Jerusalem.

#### Magdeburg

A single, high resolution, anatomical, T1-weighted scan (TR, 2500 ms; TE, 2.82 ms; TI, 1100 ms; voxel size, 1×1×1 mm3; flip angle, 7°; matrix size, 256×256×192) was acquired using a 64-channel head coil on a SIEMENS MAGNETOM Prisma 3T scanner at the University Hospital, Magdeburg, Germany.

### Data pre-processing

Surface-based morphology analysis was performed using the Freesurfer analysis suite, Version 6.0 (http://surfer.nmr.mgh.harvard.edu/). Cortical reconstruction and volumetric segmentation of the T1-weighted scans were performed automatically using the ‘recon_all’ script, described in more detail elsewhere (Dale et al., 1999; Fischl et al., 1999). In brief, the process included the removal of non-brain tissue (Ségonne et al., 2004), automated Talairach transformation, intensity normalisation (Sled et al., 1998), tessellation of the grey/white matter and pial boundaries (grey/cerebrospinal fluid) including automated topology correction and surface deformation (Dale et al., 1999; Fischl et al., 1999; Ségonne et al., 2004). After cortical models were derived, the cortical surface was inflated and registered to a sphere (Fischl et al., 1999) and the surface parcellated according to gyral and sulcal structures (Desikan et al., 2006; Fischl et al., 2004). The final surface reconstruction was inspected for potential cortical segmentation errors and, when necessary, manually corrected using the FreeView Visualisation GUI. All manually corrected reconstructions were rerun (‘autoreconall2’) utilising the edited brainmask.mgz files.

### Data analysis

A subsequent region-of-interest-based analysis was applied where we compared differences in three surface-based measures between patients and their age matched controls: mean cortical thickness (mm), surface area (mm^2^) and cortical volume (mm^3^).

Cortical thickness was measured as the shortest distance between each grey/white boundary vertex and the pial surface (white matter/cerebrospinal fluid boundary) and vice versa. The final value depicted the average of the two thickness values measured, and thickness values were then averaged across the ROI (Fischl and Dale, 2000). Surface area was measured by calculating the summed surface area across each ROI of each triangle of the surface mesh, the unit used to connect the cortical surface between each vertex. Cortical volume was computed as the sum of oblique truncated polyhedrons, as described in Winkler et al. (2018).

ROIs used for this analysis stream were derived using the anatomically defined retinotopy atlas (“benson14_retinotopy” command) implemented in the python analysis toolbox ‘neuropythy’ (Benson et al., 2014; Benson and Winawer, 2018). The atlas created several Freesurfer-based maps (visual area, eccentricity, polar angle, pRF size), which were used to delineate three ROI labels for each participant. The ROIs represented the foveal (0-2°), parafoveal (2-4°) and paracentral representations (4-8°) of V1 (Figure 1A).

Extracted values were combined across both hemispheres, where surface area and cortical volume were simply summed for each participant. For cortical thickness, the values were weighted by the respective surface area value and the mean cortical thickness value derived via following calculation:

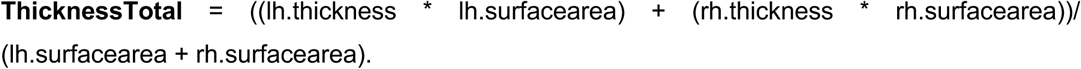

**Figure 1.**
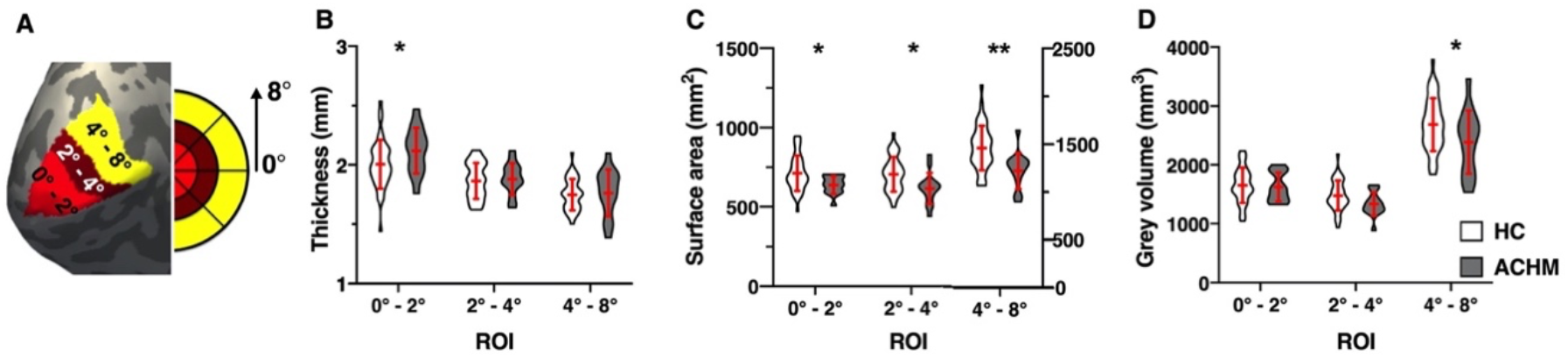
ROI surface-based morphometric values. (A) shows the left hemisphere cortical surface reconstruction of an example participant with the three overlaid ROI labels: ROI_Fovea_ (0°-2°), ROI_Parafovea_ (2°-4°) and ROI_Paracentral_ (4°-8°); violin plots show the mean cortical thickness (B), pooled surface area (C) and grey matter volume (D) in all ROIs for healthy control participants (HC, N = 42) and participants with achromatopsia (ACHM, N = 15); error bars represent +/- 1 standard deviation; *p<.05, **p<.01;

### Statistical analysis

A hierarchical linear regression was applied to predict the three outcome measures (cortical thickness, grey matter volume, surface area) in each of our three ROIs for each participant group. To account for differences in gender, age and scanner site and global brain scaling (global mean thickness, global overall surface area and estimated intracranial volume), these variables were entered in the first step of the hierarchical linear regression model (Model 1), while participant group (controls and patients) was added in the second analysis step (Model 2). All analysis steps were performed in the IBM SPSS Statistics software package, version 25. Graphs were created using Prism version 8.00 for Mac (GraphPad Software, La Jolla California USA, www.graphpad.com).

## Results

We acquired high-resolution T1-weighted images from 15 genetically confirmed participants with ACHM and compared these to 42 normally sighted individuals (Table S1) using surface-based morphometry to assess potential anatomical changes in primary visual cortex. To increase sensitivity of our analysis we used a region of interest-based approach. We identified three eccentricity bands of primary visual cortex using an anatomical atlas template (Benson et al., 2014; Benson and Winawer, 2018) which captured visual field representation in three ROIs, representing either the fovea (0-2 degrees), the parafovea (2-4 degrees) or paracentral regions of primary visual cortex (4-8 degrees) (Figure 1A).

This allowed us to determine any potential differences in cortical thickness, surface area and grey matter volume to address our specific hypotheses concerning the effect of congenital vision loss on central visual field representations of primary visual cortex. A hierarchical regression was applied to account for possible effects of age, sex, scanner site and overall brain size (global mean cortical thickness, total cortical surface area, and estimated total intracranial volume) on our more localized morphometric measures. This allowed us to determine any differences between ACHM and the control group without the influence of these potentially confounding factors (Table 1).

**Table 1.** Summary of Hierarchical Regression Analysis. Mean cortical thickness, surface area and grey matter volume were extracted from indicated ROIs and combined across hemispheres. Mean values and standard error of mean are denoted for each metric and ROI as well as F-values, degrees of freedom and associated p-values; ROI – region of interest, ACHM = Achromatopsia, HC = healthy control; Significant differences for each metric for either Model 1 (scanner site, age, gender, global brain metric) or Model 2 (participant group): * p<.05, **p<.01, ***p<.001

|  |  |  |  | Model 1 |  |  | Model 2 |  |  |
| --- | --- | --- | --- | --- | --- | --- | --- | --- | --- |
|  |  | ACHM | HC | F(5,51) | p-value |  | F(1,50) | p-value |  |
| Thickness (mm) | Fovea | 2.12 ± 0.19 | 2.007 ± 0.21 | 4.966 | .001 | ** | 6.173 | .016 | * |
|  | Parafovea | 1.88 ± 0.13 | 1.866 ± 0.15 | 5.158 | .001 | ** | 0.046 | .832 |  |
|  | Paracentral | 1.765 ± 0.20 | 1.750 ± 0.13 | 2.837 | .025 | * | 0.224 | .638 |  |
| Surface area (mm<sup>2</sup>) | Fovea | 639.00 ± 55.84 | 713.29 ± 111.39 | 14.508 | > .001 | *** | 5.651 | .021 | * |
|  | Parafovea | 616.67 ± 98.57 | 706.62 ± 232.01 | 6.194 | > .001 | *** | 4.846 | .032 | * |
|  | Paracentral | 1224.67 ± 189.24 | 1462.71 ± 35.80 | 1.633 | .168 |  | 12.851 | .001 | ** |
| Grey matter volume (mm<sup>3</sup>) | Fovea | 1623.20 ± 245.34 | 1652 ± 298.65 | 8.357 | > .001 | *** | 0.027 | .871 |  |
|  | Parafovea | 1334.07 ± 200.82 | 1478.12 ± 255.51 | 6.149 | > .001 | *** | 3.167 | .081 |  |
|  | Paracentral | 2386.87 ± 536.44 | 2686.36 ± 448.52 | 1.050 | .399 |  | 5.398 | .024 | * |

We first assessed the impact of absent cone input on cortical thickness. As shown in Figure 1B, a significant increase in cortical thickness was found in ACHM within the most foveal representation (ΔR^2^ = 7.4% ; (F (1,50) = 6.173, p = .016), while no increase in cortical thickness was seen within the parafoveal and paracentral representations of primary visual cortex in ACHM (2-4 degree: ΔR^2^ = 0.1 %; F(1,50) = 0.046, p = .832; 4-8 degree: ΔR^2^ = 0.3%; F (1,50) = 0.224, p = .638). This is consistent with the, albeit more widespread, increased cortical thickness found in congenitally blind individuals, but critically, it is localised to the foveal representation, and thus specific to the region of primary visual cortex that is deprived of visual input.

Next, we compared surface area of each ROI between groups (Figure 1C), and found that it was significantly reduced in ACHM across all ROIs, with the highest reduction found in the paracentral region (0-2 degree: ΔR2 = 4.2 %; F(1,50) = 5.651, p = .021; 2-4 degree: ΔR2 = 5.5 %; F(1,50) = 4.846, p = .032; 4-8 degree: ΔR2= 17.6%; F (1,50) =12.851, p = .001). This broader areal reduction in contrast to the more localized cortical thickening is of interest as it indicates that loss of cone input has more widespread effects on surface area.

Finally, we assessed grey matter volume. In contrast to surface area, grey matter volume was only significantly reduced paracentrally (ΔR2 = 8.8%; F (1,50) = 5.398, p = .024), as depicted in Figure 1D. In contrast, foveal and parafoveal representations of primary visual cortex showed no significant change in cortical grey matter volume (0-2 degree: ΔR2 = 0 %; F (1,50) = 0.027, p = .871; 2-4 degree: ΔR2 = 3.7 %; F (1,50) = 3.167, p = .081).

## Discussion

We applied surface-based morphometry to analyse cortical structure in ACHM, a patient population with congenitally lack of cone function, resulting in a complete loss of input from the fovea. In line with our hypotheses, we reported thicker primary visual cortex in the representations of the region of absolute loss of vision in these patients. We also found a more widespread reduction in the surface area of primary visual cortex within the central representation (0-8 degrees) we assessed.

### Cortical thickening localized to foveal visual field representations

The cortical thickening observed in the central visual field representation is consistent with that reported in several other studies of congenital total blindness and is commonly attributed to aberrant pruning processes due to absent sensory input (Aguirre et al., 2017; Bourgeois et al., 1989; Guerreiro et al., 2015; Park et al., 2009; Stryker and Harris, 1986). Increased cortical thickness has also been observed in cases of acquired localised vision loss, but in areas adjacent to the deafferented region. In this situation, it has been proposed that cortical thickening may represent a form of cortical plasticity, as areas adjacent to the representation of the lesion are likely to be used more frequently (Burge et al., 2016). Our use of localised regions of interest clearly demonstrated that increased cortical thickness was present only in the deafferented foveal representation, supporting the previously raised notion of disrupted pruning. Several studies have also demonstrated that in congenitally blind individuals, the cortical regions with increased thickness frequently process information from other sensory modalities, commonly referred to as cross-modal plasticity (Anurova et al., 2015; Bavelier and Neville, 2002; Bedny et al., 2011; Cohen et al., 1999, 1997; Cunningham et al., 2015; Guerreiro et al., 2015; Sadato et al., 2002, 1996). It is yet to be seen if the localised increase in cortical thickness found in ACHM patients is also correlated with cross-modal plasticity.

At first sight, an increase in thickness of the deafferented visual cortex might appear counterintuitive. The discrepancy in the direction of macrostructural changes has been generally attributed to different developmental trajectories of horizontal and vertical cortical dimensions (Kelly et al., 2015; Park et al., 2009; Rakic, 1995; Wierenga et al., 2014). In this respect, increased cortical thickness might be related to aberrant cortical maturation, where synaptic pruning, a process to abolish weaker cortical connections, is halted due to missing sensory input (Aguirre et al., 2017; Bourgeois et al., 1989; Guerreiro et al., 2015; Park et al., 2009; Stryker and Harris, 1986). Importantly, the standardised automated algorithm used to define cortical thickness (Fischl and Dale, 2000), is susceptible to the degree of intracortical myelination. Thus, the apparent change in cortical thickness observed may reflect differences in myelination between patients and controls (Aguirre et al., 2016; Glasser and Van Essen, 2011; Park et al., 2009). A recent study by Natu et al. (2019) indicated that the apparent thinning of the cortex over the course of normal development was correlated with an increase in myelination. Consequently, the apparent thickening observed in ACHM may reflect a failure to increase or even a decrease in cortical myelination. Our results thus lend further support to the possibility that missing sensory input can affect developmental myelination processes. Future research measuring cortical myelin in achromatopsia could shed light on this hypothesis.

### Broad reduction in surface area

A decrease in overall surface area in primary visual cortex is commonly reported in early total blindness affecting the entire visual field (Aguirre et al., 2016; Noppeney et al., 2005; Pan et al., 2007; Park et al., 2009; Ptito et al., 2008). Most studies that reported changes in early blindness focused either on the whole primary visual cortex or on the pericalcarine areas (Aguirre et al., 2016; Park et al., 2009) and included participants with blindness from a variety of causes and with a variety of visual field defects. In our ACHM cohort, surface area reduction was found across the entire extent of primary visual cortex measured (0-8deg), beyond the small, localised absolute visual field defect (Baseler et al., 2002).

Surface area is known to reach a maximum later in life, around the age of nine (Mills et al., 2014; Raznahan et al., 2011). Interestingly, pericalcarine areas do not seem to follow this general trend and no age-related peak could be observed (Wierenga et al., 2014). An earlier study reported that surface area peaks shortly after birth, especially within the highly convoluted foveal representation important for central vision (Leuba and Kraftsik, 1994). These are precisely the cortical regions that are deafferented in ACHM, potentially leading to profound effects on cortical maturation. This in turn might impact on the subsequent development of more peripheral representations, possibly explaining the broader area of reduction we observed here. Moreover, while the highest cone density is found at the foveola, leading to a central absolute scotoma in ACHM, cone photoreceptors decrease in number with eccentricity, but are still fairly numerous up to an eccentricity of 15 degrees (Curcio et al., 1991, 1990; Osterberg, 1937). Thus, the visual defect in ACHM is not limited to the foveola, supporting the possibility that the absence of cone signalling may have more widespread consequences on cortical architecture. Some recent studies also highlighted an ACHM subpopulation with compromised rod function which could be observed across all different genetic backgrounds (Khan et al., 2007; Maguire et al., 2018; Moskowitz et al., 2009; Wang et al., 2012; Zelinger et al., 2015). If this cohort also encompasses such individuals, this would also lead to a more severe reduction in surface area beyond the representation of the absolute rod scotoma.

### Reduction in grey matter volume

Although an overall decrease in grey matter volume has been reported in the occipital lobes for congenital, complete blindness (Boucard et al., 2009; Bridge et al., 2009; Burge et al., 2016; Plank et al., 2011; Prins et al., 2016b), we found only a subtle reduction in grey volume outside the retinotopic presentation of the absolute scotoma in ACHM. While grey matter volume encapsulates both cortical thickness and surface area (Winkler et al., 2018, 2010), it was shown that grey matter volume is generally more influenced by changes to surface area rather than cortical thickness (Aguirre et al., 2016; Winkler et al., 2018). This is supported by our data, where a reduction in grey matter volume was only evident in the paracentral representations of visual cortex where surface area was most reduced. Conversely, grey matter volume remained constant in the foveal representation, where surface area and thickness changed in opposite directions. This finding clearly highlights that without further metrics such as surface area and thickness, the true extent of changes to cortical structure might not be captured. This underlines the limited specificity of measurements restricted to cortical volume (Winkler et al., 2018).

### Implications of changes to cortical microstructure for cortical remapping and prospects for current restorative approaches

At this point, it is unclear to what extent cortical changes will impact current vision restoration therapies in this patient population. For example, increased cortical thickness in individuals who were born blind remained even after the cause of blindness e.g., cataracts, was reversed. Moreover, cortical thickness was negatively correlated with visual task performance while auditory task performance was positively correlated (Guerreiro et al., 2016, 2015). It appears therefore that the outcome of restoring vision can be explicitly linked to the way in which the visual cortex has been shaped during periods of deprivation. While increased cortical thickness and related cross-modal plasticity can be a limiting factor (Guerreiro et al., 2016, 2015) and are therefore likely to shape the extent of the recovery, reports of successful gene augmentation therapy in LCA suggest that cortical thickening per se is unlikely to rule out all aspects of recovery (Ashtari et al., 2015; Bennett et al., 2016).

Importantly, cortical thickness is significantly and inversely related to the age at blindness onset (Li et al., 2017). Moreover, a study by Aguirre et al. (2017) suggested that the integrity of the postretinal pathways also plays an important role in successful vision restoration, which has not yet been investigated in ACHM. While the underlying mechanisms of cortical thickening are not yet clear, the link to cross-modal plasticity and its limiting effects on therapeutic outcome are important factors to consider. Combined with the distinct reduction in surface area this suggests that early intervention will be beneficial to allow for normal cortical development in ACHM.

## Conclusion

Here, we demonstrate a specific structural alteration of the primary visual cortex in ACHM that is associated with the congenital absence of input from the cone photoreceptors. Besides a substantial reduction in cortical surface area we report a highly localized thickening of cortical areas representing the fovea. This cortical alteration is likely experience dependent and might interfere with the restoration of cone function. Consequently, our results suggest that early intervention is preferable when considering vision restoration treatment. Longitudinal studies in this patient population would help to define a time course for the observed cortical changes in order to determine the ideal time point for clinical interventions to maximise treatment efficacy.

## Data Availability

Within the limits of privacy issues of clinical data and the need for approval from the site-specific local ethics committee, data will be made available upon request. This study used openly available software and code, namely Freesurfer analysis suite, Version 6.0 (https://surfer.nmr.mgh.harvard.edu/) and the retinotopy atlas included in the python toolbox 'neuropythy' (https://github.com/noahbenson/neuropythy).

## Data availability

Within the limits of privacy issues of clinical data and the need for approval from the site-specific local ethics committee, data will be made available upon request. This study used openly available software and code, namely Freesurfer analysis suite, Version 6.0 (https://surfer.nmr.mgh.harvard.edu/) and the retinotopy atlas included in the python toolbox neuropythy’ (https://github.com/noahbenson/neuropythy).

## Author Contributions

Barbara Molz: conceptualization, data collection, data analysis, original draft, writing revised manuscript

Anne Herbik: data collection, data analysis, revised manuscript;

Heidi A. Baseler: conceptualization, resources, revised manuscript;

Pieter B. de Best: data collection, revised manuscript;

Richard Vernon: data analysis, revised manuscript;

Noa Raz: data collection, revised manuscript;

Andre Gouws: data analysis, revised manuscript;

Khazar Ahmadi: data collection, revised manuscript;

Rebecca Lowndes: data collection, revised manuscript;

Rebecca J. McLean : resources, revised manuscript;

Irene Gottlob : resources, revised manuscript;

Susanne Kohl: resources, revised manuscript;

Lars Choritz : resources, revised manuscript;

John Maguire: resources, revised manuscript;

Martin Kanowski: resources, revised manuscript;

Barbara Käsmann-Kellner: resources, revised manuscript; Ilse Wieland: resources, revised manuscript;

Eyal Banin: resources, revised manuscript;

Netta Levin: conceptualization, resources, revised manuscript, funding acquisition;

Michael B. Hoffmann: conceptualization, resources, revised manuscript, funding acquisition;

Antony B. Morland: conceptualization, resources, revised manuscript, funding acquisition;

## Acknowledgments

We thank the German patient association ‘Achromatopsie Selbsthilfe e.V.’ for support in participant recruitment. This project was supported by European Union’s Horizon 2020 research and innovation programme under the Marie Sklodowska-Curie grant agreement (No. 641805) and the German Research Foundation (DFG, HO 2002/12-1).

## Supplementary Information

### Supplementray tables

**Table S1.**
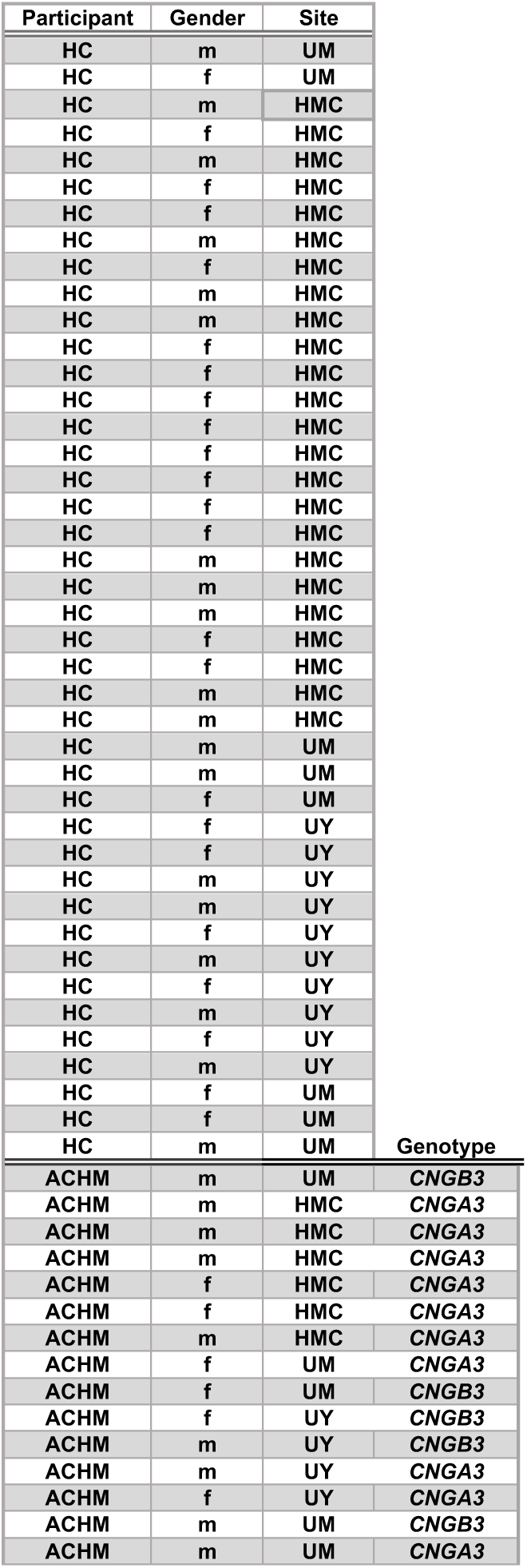
Participant demographics. Overview table summarising participant type, scanner site, age, gender, and genotype of participants with achromatopsia; (HC = healthy control, ACHM = Participant with achromatopsia; UY = University of York, UM = University of Magdeburg, HMC =Hadassah Medical Centre; m = male, f = female);

## References

Aboshiha, J., Dubis, A.M., Carroll, J., Hardcastle, A.J., Michaelides, M., 2016. The cone dysfunction syndromes. British Journal of Ophthalmology. https://doi.org/10.1136/bjophthalmol-2014-306505

Aguirre, G.K., Butt, O.H., Datta, R., Roman, A.J., Sumaroka, A., Schwartz, S.B., Cideciyan, A. V., Jacobson, S.G., 2017. Postretinal structure and function in severe congenital photoreceptor blindness caused by mutations in the GUCY2D gene. Investigative Ophthalmology and Visual Science 58, 959–973. https://doi.org/10.1167/iovs.16-20413

Aguirre, G.K., Datta, R., Benson, N.C., Prasad, S., Jacobson, S.G., Cideciyan, A. V., Bridge, H., Watkins, K.E., Butt, O.H., Dain, A.S., Brandes, L., Gennatas, E.D., 2016. Patterns of individual variation in visual pathway structure and function in the sighted and blind. PLoS ONE 11, e0164677. https://doi.org/10.1371/journal.pone.0164677

Anurova, I., Renier, L.A., Volder, A.G. De Carlson, S., Rauschecker, J.P., 2015. Relationship Between Cortical Thickness and Functional Activation in the Early Blind. Cerebral Cortex (New York, NY) 25, 2035. https://doi.org/10.1093/CERCOR/BHU009

Ashtari, M., Zhang, H., Cook, P.A., Cyckowski, L.L., Shindler, K.S., Marshall, K.A., Aravand, P., Vossough, A., Gee, J.C., Maguire, A.M., Baker, C.I., Bennett, J., 2015. Plasticity of the human visual system after retinal gene therapy in patients with Leber’s congenital amaurosis. Science translational medicine 7, 296ra110. https://doi.org/10.1126/scitranslmed.aaa8791

Baseler, H.A., Brewer, A.A., Sharpe, L.T., Morland, A.B., Jaägle, H., Wandell, B.A., 2002. Reorganization of human cortical maps caused by inherited photoreceptor abnormalities. Nature Neuroscience 5, 364– 370. https://doi.org/10.1038/nn817

Bavelier, D., Neville, H.J., 2002. Cross-modal plasticity: where and how? Nature Reviews Neuroscience 3, 443–452. https://doi.org/10.1038/nrn848

Bedny, M., Pascual-Leone, A., Dodell-Feder, D., Fedorenko, E., Saxe, R., 2011. Language processing in the occipital cortex of congenitally blind adults. Proceedings of the National Academy of Sciences 108, 4429–4434. https://doi.org/10.1073/pnas.1014818108

Bennett, J., Wellman, J., Marshall, K.A., McCague, S., Ashtari, M., DiStefano-Pappas, J., Elci, O.U., Chung, D.C., Sun, J., Wright, J.F., Cross, D.R., Aravand, P., Cyckowski, L.L., Bennicelli, J.L., Mingozzi, F., Auricchio, A., Pierce, E.A., Ruggiero, J., Leroy, B.P., Simonelli, F., High, K.A., Maguire, A.M., 2016. Safety and durability of effect of contralateral-eye administration of AAV2 gene therapy in patients with childhood-onset blindness caused by RPE65 mutations: a follow-on phase 1 trial. The Lancet 388, 661–672. https://doi.org/10.1016/S0140-6736(16)30371-3

Benson, N.C., Butt, O.H., Brainard, D.H., Aguirre, G.K., 2014. Correction of Distortion in Flattened Representations of the Cortical Surface Allows Prediction of V1-V3 Functional Organization from Anatomy. PLoS Computational Biology 10, e1003538. https://doi.org/10.1371/journal.pcbi.1003538

Benson, N.C., Winawer, J., 2018. Bayesian analysis of retinotopic maps. eLife 7. https://doi.org/10.7554/eLife.40224

Boucard, C.C., Hernowo, A.T., Maguire, R.P., Jansonius, N.M., Roerdink, J.B.T.M., Hooymans, J.M.M., Cornelissen, F.W., 2009. Changes in cortical grey matter density associated with long-standing retinal visual field defects. Brain 132, 1898–1906. https://doi.org/10.1093/brain/awp119

Bourgeois, J.P., Jastreboff, P.J., Rakic, P., 1989. Synaptogenesis in visual cortex of normal and preterm monkeys: evidence for intrinsic regulation of synaptic overproduction. Proceedings of the National Academy of Sciences 86, 4297–4301. https://doi.org/10.1073/pnas.86.11.4297

Bridge, H., Cowey, A., Ragge, N., Watkins, K.E., 2009. Imaging studies in congenital anophthalmia reveal preservation of brain architecture in “visual” cortex. Brain 132, 3467–3480. https://doi.org/10.1093/brain/awp279

Bridge, H., von dem Hagen, E.A.H., Davies, G., Chambers, C., Gouws, A.D., Hoffmann, M.B., Morland, A.B., 2014. Changes in brain morphology in albinism reflect reduced visual acuity. Cortex 56, 64–72. https://doi.org/10.1016/j.cortex.2012.08.010

Burge, W.K., Griffis, J.C., Nenert, R., Elkhetali, A., Decarlo, D.K., Ver Hoef, L.W., Ross, L.A., Visscher, K.M., 2016. Cortical thickness in human V1 associated with central vision loss. Scientific Reports 6, 23268. https://doi.org/10.1038/srep23268

Cohen, L.G., Celnik, P., Pascual-Leone, A., Corwell, B., Faiz, L., Dambrosia, J., Honda, M., Sadato, N., Gerloff, C., Dolores Catalá, M., Hallett, M., 1997. Functional relevance of cross-modal plasticity in blind humans. Nature 389, 180–183. https://doi.org/10.1038/38278

Cohen, L.G., Weeks, R.A., Sadato, N., Celnik, P., Ishii, K., Hallett, M., 1999. Period of susceptibility for cross-modal plasticity in the blind. Annals of neurology 45, 451–60.

Cunningham, S.I., Weiland, J.D., Bao, P., Lopez-Jaime, G.R., Tjan, B.S., 2015. Correlation of vision loss with tactile-evoked V1 responses in retinitis pigmentosa. Vision Research 111, 197–207. https://doi.org/10.1016/j.visres.2014.10.015

Curcio, C.A., Allen, K.A., Sloan, K.R., Lerea, C.L., Hurley, J.B., Klock, I.B., Milam, A.H., 1991. Distribution and morphology of human cone photoreceptors stained with anti-blue opsin. Journal of Comparative Neurology 312, 610–624. https://doi.org/10.1002/cne.903120411

Curcio, C.A., Sloan, K.R., Kalina, R.E., Hendrickson, A.E., 1990. Human photoreceptor topography. Journal of Comparative Neurology 292, 497–523. https://doi.org/10.1002/cne.902920402

d’Almeida, O.C., Mateus, C., Reis, A., Grazina, M.M., Castelo-Branco, M., 2013. Long term cortical plasticity in visual retinotopic areas in humans with silent retinal ganglion cell loss. NeuroImage 81, 222–230. https://doi.org/10.1016/j.neuroimage.2013.05.032

Dale, A.M., Fischl, B., Sereno, M.I., 1999. Cortical surface-based analysis: I. Segmentation and surface reconstruction. NeuroImage 9, 179–194. https://doi.org/10.1006/nimg.1998.0395

Desikan, R.S., Ségonne, F., Fischl, B., Quinn, B.T., Dickerson, B.C., Blacker, D., Buckner, R.L., Dale, A.M., Maguire, R.P., Hyman, B.T., Albert, M.S., Killiany, R.J., 2006. An automated labeling system for subdividing the human cerebral cortex on MRI scans into gyral based regions of interest. NeuroImage 31, 968–80. https://doi.org/10.1016/j.neuroimage.2006.01.021

Fischer, M.D., Michalakis, S., Wilhelm, B., Zobor, D., Muehlfriedel, R., Kohl, S., Weisschuh, N., Ochakovski, G.A., Klein, R., Schoen, C., Sothilingam, V., Garcia-Garrido, M., Kuehlewein, L., Kahle, N., Werner, A., Dauletbekov, D., Paquet-Durand, F., Tsang, S., Martus, P., Peters, T., Seeliger, M., Bartz-Schmidt, K.U., Ueffing, M., Zrenner, E., Biel, M., Wissinger, B., 2020. Safety and Vision Outcomes of Subretinal Gene Therapy Targeting Cone Photoreceptors in Achromatopsia: A Nonrandomized Controlled Trial. JAMA Ophthalmology 138, 643–651. https://doi.org/10.1001/jamaophthalmol.2020.1032

Fischl, B., Dale, A.M., 2000. Measuring the thickness of the human cerebral cortex from magnetic resonance images. Proceedings of the National Academy of Sciences of the United States of America 97, 11050–5. https://doi.org/10.1073/pnas.200033797

Fischl, B., Sereno, M.I., Dale, A.M., 1999. Cortical surface-based analysis: II. Inflation, flattening, and a surface-based coordinate system. NeuroImage 9, 195–207. https://doi.org/10.1006/nimg.1998.0396

Fischl, B., Van Der Kouwe, A., Destrieux, C., Halgren, E., Ségonne, F., Salat, D.H., Busa, E., Seidman, L.J., Goldstein, J., Kennedy, D., Caviness, V., Makris, N., Rosen, B., Dale, A.M., 2004. Automatically Parcellating the Human Cerebral Cortex. Cerebral Cortex 14, 11–22. https://doi.org/10.1093/cercor/bhg087

Glasser, M.F., Van Essen, D.C., 2011. Mapping Human Cortical Areas In Vivo Based on Myelin Content as Revealed by T1- and T2-Weighted MRI. Journal of Neuroscience 31, 11597–11616. https://doi.org/10.1523/jneurosci.2180-11.2011

Guerreiro, M.J.S., Erfort, M. V., Henssler, J., Putzar, L., Röder, B., 2015. Increased visual cortical thickness in sight-recovery individuals. Human Brain Mapping 36, 5265–5274. https://doi.org/10.1002/hbm.23009

Guerreiro, M.J.S., Putzar, L., Röder, B., 2016. Persisting Cross-Modal Changes in Sight-Recovery Individuals Modulate Visual Perception. Current Biology 26, 3096–3100. https://doi.org/10.1016/j.cub.2016.08.069

Haegerstrom-Portnoy, G., Schneck, M.E., Verdon, W.A., Hewlett, S.E., 1996. Clinical vision characteristics of the congenital achromatopsias. II. Color vision. Optometry and Vision Science 73, 457–465. https://doi.org/10.1097/00006324-199607000-00002

Hirji, N., Georgiou, M., Kalitzeos, A., Bainbridge, J.W., Kumaran, N., Aboshiha, J., Carroll, J., Michaelides, M., 2018. Longitudinal assessment of retinal structure in achromatopsia patients with long-term follow-up. Investigative Ophthalmology and Visual Science 59, 5735–5744. https://doi.org/10.1167/iovs.18-25452

Jiang, J., Zhu, W., Shi, F., Liu, Y., Li, J., Qin, W., Li, K., Yu, C., Jiang, T., 2009. Thick Visual Cortex in the Early Blind. Journal of Neuroscience 29, 2205–2211. https://doi.org/10.1523/JNEUROSCI.5451-08.2009

Kelly, K.R., Desimone, K.D., Gallie, B.L., Steeves, J.K.E., 2015. Increased cortical surface area and gyrification following long-term survival from early monocular enucleation. NeuroImage: Clinical 7, 297–305. https://doi.org/10.1016/j.nicl.2014.11.020

Khan, N.W., Wissinger, B., Kohl, S., Sieving, P.A., 2007. CNGB3 achromatopsia with progressive loss of residual cone function and impaired rod-mediated function. Investigative Ophthalmology and Visual Science 48, 3864–3871. https://doi.org/10.1167/iovs.06-1521

Lemos, J., Pereira, D., Castelo-Branco, M., 2016. Visual Cortex Plasticity Following Peripheral Damage To The Visual System: fMRI Evidence. Current Neurology and Neuroscience Reports. https://doi.org/10.1007/s11910-016-0691-0

Leuba, G., Kraftsik, R., 1994. Changes in volume, surface estimate, three-dimensional shape and total number of neurons of the human primary visual cortex from midgestation until old age. Anatomy and Embryology 190, 351–366. https://doi.org/10.1007/BF00187293

Li, Q., Song, M., Xu, J., Qin, W., Yu, C., Jiang, T., 2017. Cortical thickness development of human primary visual cortex related to the age of blindness onset. Brain Imaging and Behavior 11, 1029–1036. https://doi.org/10.1007/s11682-016-9576-8

Maguire, J., McKibbin, M., Khan, K., Kohl, S., Ali, M., McKeefry, D.J., 2018. CNGB3 mutations cause severe rod dysfunction. Ophthalmic Genetics 39, 108–114. https://doi.org/10.1080/13816810.2017.1368087

Michalakis, S., Schön, C., Becirovic, E., Biel, M., 2017. Gene therapy for achromatopsia. Journal of Gene Medicine. https://doi.org/10.1002/jgm.2944

Mills, K.L., Lalonde, F., Clasen, L.S., Giedd, J.N., Blakemore, S.J., 2014. Developmental changes in the structure of the social brain in late childhood and adolescence. Social Cognitive and Affective Neuroscience 9, 123–131. https://doi.org/10.1093/scan/nss113

Moskowitz, A., Hansen, R.M., Akula, J.D., Eklund, S.E., Fulton, A.B., 2009. Rod and rod-driven function in achromatopsia and blue cone monochromatism. Investigative Ophthalmology and Visual Science 50, 950–958. https://doi.org/10.1167/iovs.08-2544

Natu, V.S., Gomez, J., Barnett, M., Jeska, B., Kirilina, E., Jaeger, C., Zhen, Z., Cox, S., Weiner, K.S., Weiskopf, N., Grill-Spector, K., 2019. Apparent thinning of human visual cortex during childhood is associated with myelination. Proceedings of the National Academy of Sciences of the United States of America 116, 20750–20759. https://doi.org/10.1073/pnas.1904931116

Noppeney, U., Friston, K.J., Ashburner, J., Frackowiak, R., Price, C.J., 2005. Early visual deprivation induces structural plasticity in gray and white matter [1]. Current Biology. https://doi.org/10.1016/j.cub.2005.06.053

Osterberg, G., 1937. Topography of the Layer of Rods and Cones in the Human Retina. Journal of the American Medical Association 108, 232. https://doi.org/10.1001/jama.1937.02780030070033

Pan, W.J., Wu, G., Li, C.X., Lin, F., Sun, J., Lei, H., 2007. Progressive atrophy in the optic pathway and visual cortex of early blind Chinese adults: A voxel-based morphometry magnetic resonance imaging study. NeuroImage 37, 212–220. https://doi.org/10.1016/j.neuroimage.2007.05.014

Park, H.J., Lee, J.D., Kim, E.Y., Park, B., Oh, M.K., Lee, S.C., Kim, J.J., 2009. Morphological alterations in the congenital blind based on the analysis of cortical thickness and surface area. NeuroImage 47, 98– 106. https://doi.org/10.1016/j.neuroimage.2009.03.076

Plank, T., Frolo, J., Brandl-Rühle, S., Renner, A.B., Hufendiek, K., Helbig, H., Greenlee, M.W., 2011. Gray matter alterations in visual cortex of patients with loss of central vision due to hereditary retinal dystrophies. NeuroImage 56, 1556–1565. https://doi.org/10.1016/j.neuroimage.2011.02.055

Prins, D., Hanekamp, S., Cornelissen, F.W., 2016a. Structural brain MRI studies in eye diseases: Are they clinically relevant? A review of current findings. Acta Ophthalmologica. https://doi.org/10.1111/aos.12825

Prins, D., Plank, T., Baseler, H.A., Gouws, A.D., Beer, A., Morland, A.B., Greenlee, M.W., Cornelissen, F.W., 2016b. Surface-Based Analyses of Anatomical Properties of the Visual Cortex in Macular Degeneration. PLOS ONE 11, e0146684. https://doi.org/10.1371/journal.pone.0146684

Ptito, M., Schneider, F.C.G., Paulson, O.B., Kupers, R., 2008. Alterations of the visual pathways in congenital blindness. Experimental Brain Research 187, 41–49. https://doi.org/10.1007/s00221-008-1273-4

Rakic, P., 1995. A small step for the cell, a giant leap for mankind: a hypothesis of neocortical expansion during evolution. Trends in Neurosciences 18, 383–388. https://doi.org/10.1016/0166-2236(95)93934-P

Raznahan, A., Shaw, P., Lalonde, F., Stockman, M., Wallace, G.L., Greenstein, D., Clasen, L., Gogtay, N., Giedd, J.N., 2011. How does your cortex grow? Journal of Neuroscience 31, 7174–7177. https://doi.org/10.1523/JNEUROSCI.0054-11.2011

Remmer, M.H., Rastogi, N., Ranka, M.P., Ceisler, E.J., 2015. Achromatopsia: A review. Current Opinion in Ophthalmology. https://doi.org/10.1097/ICU.0000000000000189

Sadato, N., Okada, T., Honda, M., Yonekura, Y., 2002. Critical period for cross-modal plasticity in blind humans: A functional MRI study. NeuroImage 16, 389–400. https://doi.org/10.1006/nimg.2002.1111

Sadato, N., Pascual-Leone, A., Grafman, J., Ibañez, V., Deiber, M.P., Dold, G., Hallett, M., 1996. Activation of the primary visual cortex by Braille reading in blind subjects. Nature 380, 526–528. https://doi.org/10.1038/380526a0

Ségonne, F., Dale, A.M., Busa, E., Glessner, M., Salat, D.H., Hahn, H.K., Fischl, B., 2004. A hybrid approach to the skull stripping problem in MRI. NeuroImage 22, 1060–1075. https://doi.org/10.1016/j.neuroimage.2004.03.032

Sled, J.G., Zijdenbos, A.P., Evans, A.C., 1998. A nonparametric method for automatic correction of intensity nonuniformity in MRI data. IEEE transactions on medical imaging 17, 87–97. https://doi.org/10.1109/42.668698

Stryker, M.P., Harris, W.A., 1986. Binocular impulse blockade prevents the formation of ocular dominance columns in cat visual cortex. The Journal of neuroscience : the official journal of the Society for Neuroscience 6, 2117–33.

Von Dem Hagen, E.A.H., Houston, G.C., Hoffmann, M.B., Jeffery, G., Morland, A.B., 2005. Retinal abnormalities in human albinism translate into a reduction of grey matter in the occipital cortex. European Journal of Neuroscience 22, 2475–2480. https://doi.org/10.1111/j.1460-9568.2005.04433.x

Wang, I., Khan, N.W., Branham, K., Wissinger, B., Kohl, S., Heckenlively, J.R., 2012. Establishing baseline rod electroretinogram values in achromatopsia and cone dystrophy. Documenta Ophthalmologica 125, 229–233. https://doi.org/10.1007/s10633-012-9350-1

Wierenga, L.M., Langen, M., Oranje, B., Durston, S., 2014. Unique developmental trajectories of cortical thickness and surface area. NeuroImage 87, 120–126. https://doi.org/10.1016/j.neuroimage.2013.11.010

Winkler, A.M., Greve, D.N., Bjuland, K.J., Nichols, T.E., Sabuncu, M.R., Håberg, A.K., Skranes, J., Rimol, L.M., 2018. Joint Analysis of Cortical Area and Thickness as a Replacement for the Analysis of the Volume of the Cerebral Cortex. Cerebral Cortex 28, 738–749. https://doi.org/10.1093/cercor/bhx308

Winkler, A.M., Kochunov, P., Blangero, J., Almasy, L., Zilles, K., Fox, P.T., Duggirala, R., Glahn, D.C., 2010. Cortical thickness or grey matter volume? The importance of selecting the phenotype for imaging genetics studies. NeuroImage 53, 1135–1146. https://doi.org/10.1016/j.neuroimage.2009.12.028

Zelinger, L., Cideciyan, A. V., Kohl, S., Schwartz, S.B., Rosenmann, A., Eli, D., Sumaroka, A., Roman, A.J., Luo, X., Brown, C., Rosin, B., Blumenfeld, A., Wissinger, B., Jacobson, S.G., Banin, E., Sharon, D., 2015. Genetics and disease expression in the CNGA3 form of achromatopsia: Steps on the path to gene therapy. Ophthalmology 122, 997–1007. https://doi.org/10.1016/j.ophtha.2014.11.025

Zobor, D., Zobor, G., Kohl, S., 2015. Achromatopsia: On the Doorstep of a Possible Therapy. Ophthalmic Research. https://doi.org/10.1159/000435957

